## Supplementary Material for "How Users of Different Ages Rate a Mobile App for Self-Testing of Hearing"

**Observation sheet**

1. **Headphones used and calibration method**

□ Headphones from the set (no calibration)

□ Other headphones (biological calibration)

□ Demonstration test (calibration omitted)

2. **Participant's independence in carrying out the test**

□ A fully self-performed test (independently)

□ Assistance at certain elements of the procedure (partial assistance):

□ Installation

□ Need to clarify the operation/rules of the test

□ Saving of the result (PDF or screenshot)

□ Sending the result to the e-mail address

□ Other

□ Testing entirely by another person (full assistance)

3. **Interpretation of the test result** (correct evaluation of the result - correct/incorrect; distinction between left and right ear; correct understanding of the X axes-frequency and Y axes- sound level)

□ Correct (all elements correct)

□ Partially correct:

□ Incorrect evaluation of the result - correct/incorrect

□ Incorrect distinction between left and right ear

□ Incorrect understanding of the X axes-frequency

□ Incorrect understanding of the Y axes-sound level

□ Other

□ Incorrect (all elements incorrect)

**Instruction: how to take a hearing test using the mobile app**

**Step 1 Preparation for the test**

To test your hearing with the app, you need an Android-based mobile device (smartphone, tablet) and headphones. You will also need internet access.

If you do not have the required equipment notify the person conducting the test - you will be provided with the appropriate equipment for the test.

**Step 2 Downloading the app**

1. Using the “Google Play” platform, install the “Hearing test” app

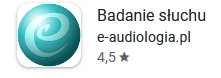

2. When the app opens, select “Hearing threshold test” and follow the instructions on the device.
3. After performing the test, record the result obtained.

**Step 3 Sending of the test results obtained**

1. Once you have carried out the tests on the app, send the results to the e-mail address below:

**……………………**

2. In the subject line of your message, write the serial number that the researcher gave you at the beginning of the meeting (e.g. 001)

Supplementary Material

**Part I**

**Table 1*.*** Questions used for rating the app, based on MARS criteria

| **Engagement–interest** | To what extent did you find the tested app interesting? | Boring, the app was not interesting |
| --- | --- | --- |
|  |  | Mostly uninteresting |
|  |  | OK, neither interesting nor uninteresting |
|  |  | Moderately interesting |
|  |  | Very interesting |
| **Engagement– customization** | Are the settings included in the app, preferences for app features (e.g. font size, notifications, etc.) satisfactory? | The app does not allow any customization of preferred features |
|  |  | For the most part, the options for preferences is insufficient |
|  |  | For the most part, the ability to change preferences is sufficient |
|  |  | Allows multiple customization options |
|  |  | Allows full customization |
| **Aesthetics** | To what extent was the app visually appealing? | Lack of visual appeal, unpleasant appearance, graphics look amateurish |
|  |  | Little visual appeal, visually boring, low quality graphics |
|  |  | Average visual appeal, graphics of moderate quality |
|  |  | High visual appeal, good quality graphics |
|  |  | Very high visual appeal, graphics professionally designed |
| **Functionality** | To what extent is the application easy to use? To what extent are the menu icons and instructions clear? | Difficult, limited instructions, menu icons are confusing and complicated |
|  |  | Suitable for use after a long time of learning to operate |
|  |  | Suitable for use after some time learning how to use it |
|  |  | Easy to learn how to use the app, instructions are clear and easy to read |
|  |  | Able to use app immediately; the system is intuitive, simple |
| **Information–quality** | Does the way feedback is presented make the test result clear and understandable? | The result obtained is completely unclear/ unintelligible |
|  |  | The result obtained is mostly unclear, missing a lot of relevant information |
|  |  | The result obtained is acceptable, but some information can be unclear |
|  |  | The result obtained is mostly clear, with minor problems in understanding some information |
|  |  | The result obtained is completely clear, there were no problems with understanding it |
| **Information–quantity** | Does the way feedback is presented make the result of the test clear and understandable? | No, the amount of information is minimal and confusing |
|  |  | No, the amount of information is insufficient and it can be confusing |
|  |  | OK, but not comprehensive or concise |
|  |  | Yes, mostly a wide range of information and no unnecessary details |
|  |  | Yes, the information is presented in both a comprehensive and concise manner |

**Part II**

**Table 2*.*** Questions used for assessing other aspects of usability

| **Difficulties in using the app** | Indicate which element of the app was the most difficult for you | Installing the app |
| --- | --- | --- |
|  |  | Device calibration |
|  |  | Performing the test |
|  |  | Saving the test result |
|  |  | Interpretation of the result |
| **Understanding the results provided by the app** | Which way of presenting the test result is the most accessible/understandable? | Audiogram (graph) |
|  |  | Description |
|  |  | Calculated degree of hearing loss |
|  |  | Other |
| **Preferred method of hearing assessment** | Which hearing test method do you prefer? | Using the app |
|  |  | Conventional test |
|  |  | No preference |
|  | If you could choose between making an appointment for a hearing test using the app at your primary care physician's office or taking a separate visit to an audiology clinic, which would you prefer? | Test using the app at my primary care physician's office |
|  |  | Conventional test in an audiology clinic |
